# QUEST-AI: A System for Question Generation, Verification, and Refinement using AI for USMLE-Style Exams

**DOI:** 10.1101/2023.04.25.23288588

**Authors:** Suhana Bedi, Scott L. Fleming, Chia-Chun Chiang, Keith Morse, Aswathi Kumar, Birju Patel, Jenelle A. Jindal, Conor Davenport, Craig Yamaguchi, Nigam H. Shah

**Affiliations:** Department of Biomedical Data Science, Stanford University, Stanford, CA, USA; SmarterDx, Inc. Brooklyn, NY, USA; Department of Neurology, Mayo Clinic, Rochester, MN; Department of Pediatrics, Stanford School of Medicine, Stanford, CA, USA; Department of Medicine, Stanford School of Medicine, Stanford, CA, USA; Center for Biomedical Informatics Research, Stanford, CA, USA; Western University of Health Sciences, Pomona, California, USA; Human-Centered Artificial Intelligence Institute, Stanford University, Stanford, CA, USA; Clinical Excellence Research Center, Stanford School of Medicine, Stanford, CA, USA; Technology and Digital Solutions, Stanford Health Care, Palo Alto, California, USA

**Keywords:** USMLE, Medical Education, Large Language Models, Artificial Intelligence, GPT-4, Exam Question Generation, Automated Assessment, Medical Exam Preparation, Question Validity, Medical Licensing Examination

## Abstract

The United States Medical Licensing Examination (USMLE) is a critical step in assessing the competence of future physicians, yet the process of creating exam questions and study materials is both time-consuming and costly. While Large Language Models (LLMs), such as OpenAI’s GPT-4, have demonstrated proficiency in answering medical exam questions, their potential in generating such questions remains underexplored. This study presents QUEST-AI, a novel system that utilizes LLMs to (1) generate USMLE-style questions, (2) identify and flag incorrect questions, and (3) correct errors in the flagged questions. We evaluated this system’s output by constructing a test set of 50 LLM-generated questions mixed with 50 human-generated questions and conducting a two-part assessment with three physicians and two medical students. The assessors attempted to distinguish between LLM and human-generated questions and evaluated the validity of the LLM-generated content. A majority of exam questions generated by QUEST-AI were deemed valid by a panel of three clinicians, with strong correlations between performance on LLM-generated and human-generated questions. This pioneering application of LLMs in medical education could significantly increase the ease and efficiency of developing USMLE-style medical exam content, offering a cost-effective and accessible alternative for exam preparation.

## 1. Introduction

Every year, over 100,000 medical students take the United States Medical Licensing Examination (USMLE), administered by the National Board of Medical Examiners (NBME).^1^ This rigorous examination is crucial for ensuring the competence of future physicians. However, generating the exam questions and related preparation materials is a manual process, which is both time-consuming and costly. On average, each student spends over $4,000 on buying USMLE-related study materials.^2^ The high costs and substantial effort associated with producing these materials are the primary drivers of the cost, and offer a great opportunity for technological intervention.

The quality of these exam questions plays a critical role in medical education and the training of future healthcare professionals. These exams assess key clinical knowledge and decision-making skills, which directly influence how prepared medical students are to handle real-world patient care. Ensuring the accuracy and biological relevance of the questions is vital for maintaining high standards in healthcare, as the competence of future physicians ultimately impacts patient outcomes.

The adoption of Artificial Intelligence (AI) in healthcare is rapidly increasing, driven by advancements in Generative AI and especially, Large Language Models (LLMs) such as OpenAI’s GPT-4.^3,4,5^ LLMs have been explored for various use cases in medicine, including generating clinical notes, summarizing patient records, and providing decision support.^6,7,8^ Numerous studies have demonstrated the proficiency of these models in answering USMLE questions, achieving over 80% accuracy on the USMLE Step 2 Clinical Knowledge (CK) exam.^9^ Despite their success in answering exam questions, there is limited research on the use of LLMs for *generating* medical exam questions, particularly for the USMLE. To address this gap, we introduce QUEST-AI, an autonomous system powered by LLMs that (1) generates USMLE-style questions based on in-context examples, (2) verifies the system-generated questions using an ensemble of LLMs, and (3) refines any questions identified as incorrect. The system is evaluated with the assistance of physicians and medical students.

We began by prompting GPT-4 to generate 50 questions inspired by sample questions from the USMLE Step 2 Clinical Knowledge (CK) exam. Then, we used aggregated predictions from an ensemble of diverse LLMs to flag incorrect questions. Finally, we prompted GPT-4 again to correct the flawed questions. In order to evaluate the quality of questions generated using our approach, we constructed a test set containing our 50 system-generated questions randomly interspersed with 50 human-generated sample questions. Three physicians and two medical students engaged in a twofold assessment: (1) they attempted to distinguish between the system-generated and human-generated USMLE-style questions, and (2) they assessed the validity of the system-generated questions and answers.

To our knowledge, ours is the first study to generate, verify, and refine USMLE-style questions using LLMs (Figure 1). This shift from answering questions to generating questions represents a novel application of AI in medical education, with the potential to revolutionize exam content development.

**Figure 1:**
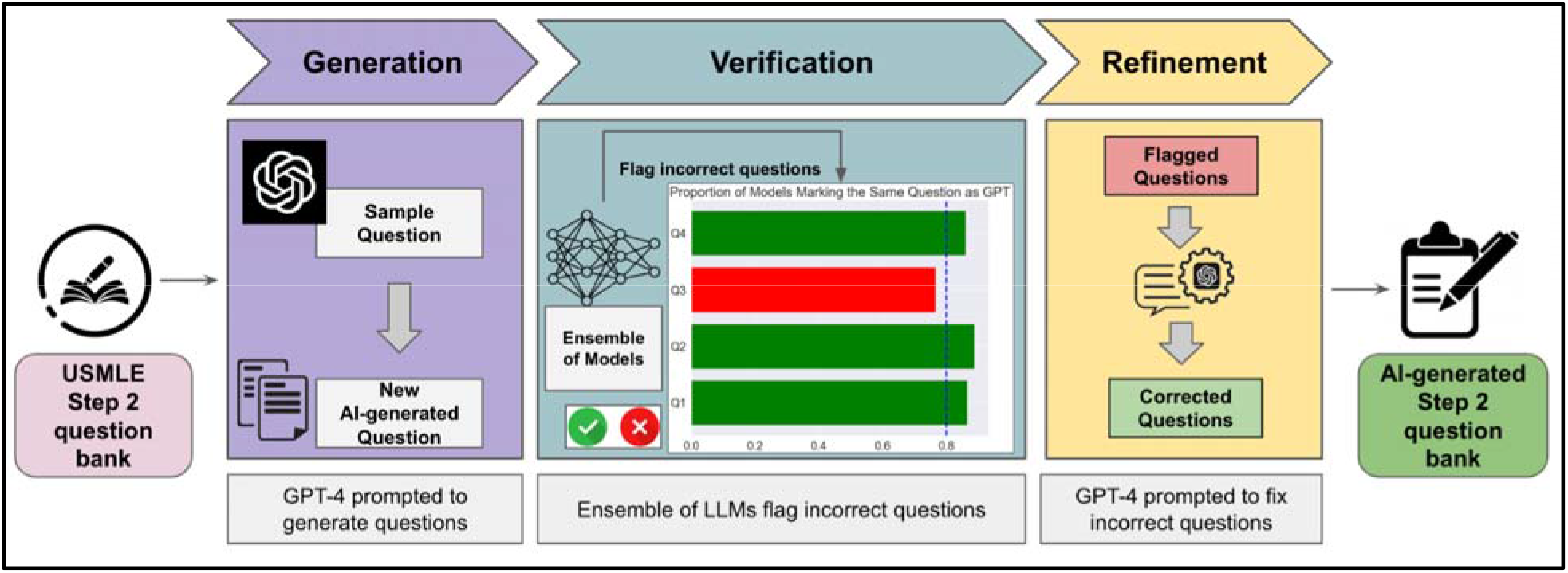
The QUEST-AI System for Generation, Verification, and Refinement of USMLE-Style Questions: This figure illustrates the process used by QUEST-AI to generate, verify, and refine USMLE-style questions. The process begins with GPT-4 generating questions using sample questions from the USMLE Step 2 question bank as in-context examples. An ensemble of LLMs then processes these questions, flagging any incorrect ones based on their ensembled predictions. Finally, GPT-4 refines the flagged questions, resulting in a high-quality, system-generated Step 2 question bank.

## 2. Related Work

The use of Large Language Models (LLMs) in healthcare and education has seen considerable growth and innovation.

### 2.1. LLMs in Healthcare

LLMs have become prominent in healthcare due to their advanced natural language processing capabilities, allowing them to handle large datasets and generate accurate, contextually relevant text.^10^ Bedi et al^11^ provide a systematic review of LLM applications across various healthcare tasks, including diagnosis^12^, report generation^13^, treatment recommendations^14^, and clinical referrals.^14,15^ While these studies demonstrate the potential of LLMs in clinical settings, few have explored their application in the educational domain, specifically for training future healthcare professionals. This study builds on these advancements, applying LLMs to a novel task: automated medical exam question generation.

### 2.2. LLMs in Education

LLMs have shown great promise in education, particularly in providing real-time support and feedback across a range of subjects, such as math ^16^, law ^17^ and medicine ^4^.

Recent research has applied LLMs for automatic question generation, improving educational content and assessment quality. Laverghetta Jr. and Licato demonstrated the use of GPT-3 for cognitive assessments, and Tran et al. applied GPT-4 to generate high-quality multiple-choice questions (MCQs) for computing courses. ^18 19^ However, these efforts focus on general education, with little attention given to specialized medical education, particularly for high-stakes exams like the USMLE.

### 2.3. LLMs in Medical Education

There has been growing interest in using LLMs to generate medical exam questions due to their potential to reduce the burden on educators and streamline content creation. A systematic review by Artsi et al. discovered a total of eight studies that explored LLMs like GPT-3.5 and GPT-4 for producing valid multiple-choice questions (MCQs) across various medical disciplines, including neurosurgery, internal medicine, and dermatology.^20^ While these studies demonstrate the feasibility of LLMs in medical education, they also highlight limitations such as inaccuracies, lower complexity in generated questions, and a lack of rigorous evaluation of content quality and validity, particularly for high-stakes exams like the USMLE. ^21 22 23 24 25^

To address these gaps, our study evaluates GPT-4’s ability to generate USMLE Step 2 CK-style exam questions. We provide insights into the practical applications of AI in medical education and its potential to enhance the accessibility and quality of exam preparation materials. By presenting a fully autonomous system for generating, verifying, and refining USMLE-style questions, we aim to demonstrate the capacity of LLMs to generate high-quality exam content, thereby improving the development and accessibility of medical education resources.

## 3. Methods

### 3.1. Data collection and generation

We randomly selected a set of 50 human-generated questions from a bank of 120 publicly available USMLE Step 2 CK test sample questions, ensuring that these questions did not include associated images or abstracts^26^. This was done to maintain a controlled and uniform format for comparison purposes.

For system-generated questions, we employed a prompt chaining approach with GPT-4 as shown in Figure 2. We started with a human-generated USMLE CK test question-answer pair, which was included in the initial prompt to GPT-4. The model then generated an explanation of why the given answer was correct and the others were incorrect. This original question, along with the system-generated explanation, were used in a follow-up prompt instructing GPT-4 to generate another USMLE Step 2 CK-style question in a similar format. This method ensured the generated questions closely matched the format, style, and complexity of the human-generated ones, promoting consistency and reducing deviations from the desired standards.

**Figure 2:**
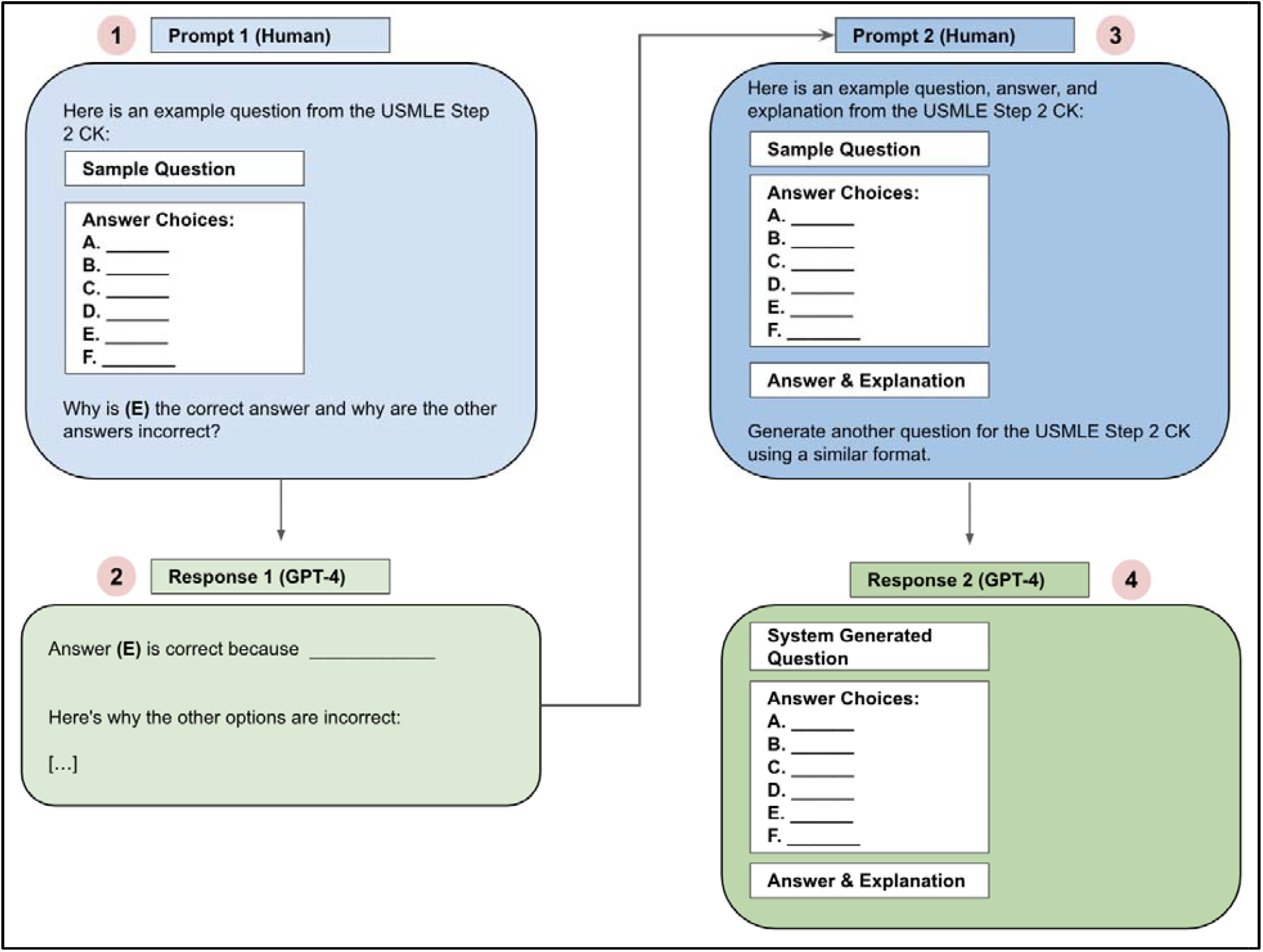
Prompt chaining strategy for question generation: First, we provide GPT-4 with an example question from the USMLE CK exam and ask why a specific option is correct and why others are incorrect. Once GPT-4 generates a response, we create a new prompt incorporating this response and the original question, then ask GPT-4 to generate another question in a similar format.

After generating a set of system-generated questions, we compiled these alongside the human-generated ones and randomly shuffled them to create a comprehensive 100-question set. This randomization was crucial to ensure an unbiased evaluation.

### 3.2. Evaluation by Physicians

A group of three licensed, practicing physicians and two medical students were tasked with evaluating the 100-question set. They were instructed to:

1. Choose the single best answer to each question without consulting any external reference.

2. Guess whether each question was generated by humans or GPT-4.

In a separate task, three physicians reviewed the 50 system-generated exam questions to evaluate their correctness, using any available external references. They recorded the type of errors found in the system-generated questions and the time taken to make their determinations. The two phases of the study, marked by the different tasks performed by the medical specialists, are illustrated in Figure 3.

**Figure 3:**
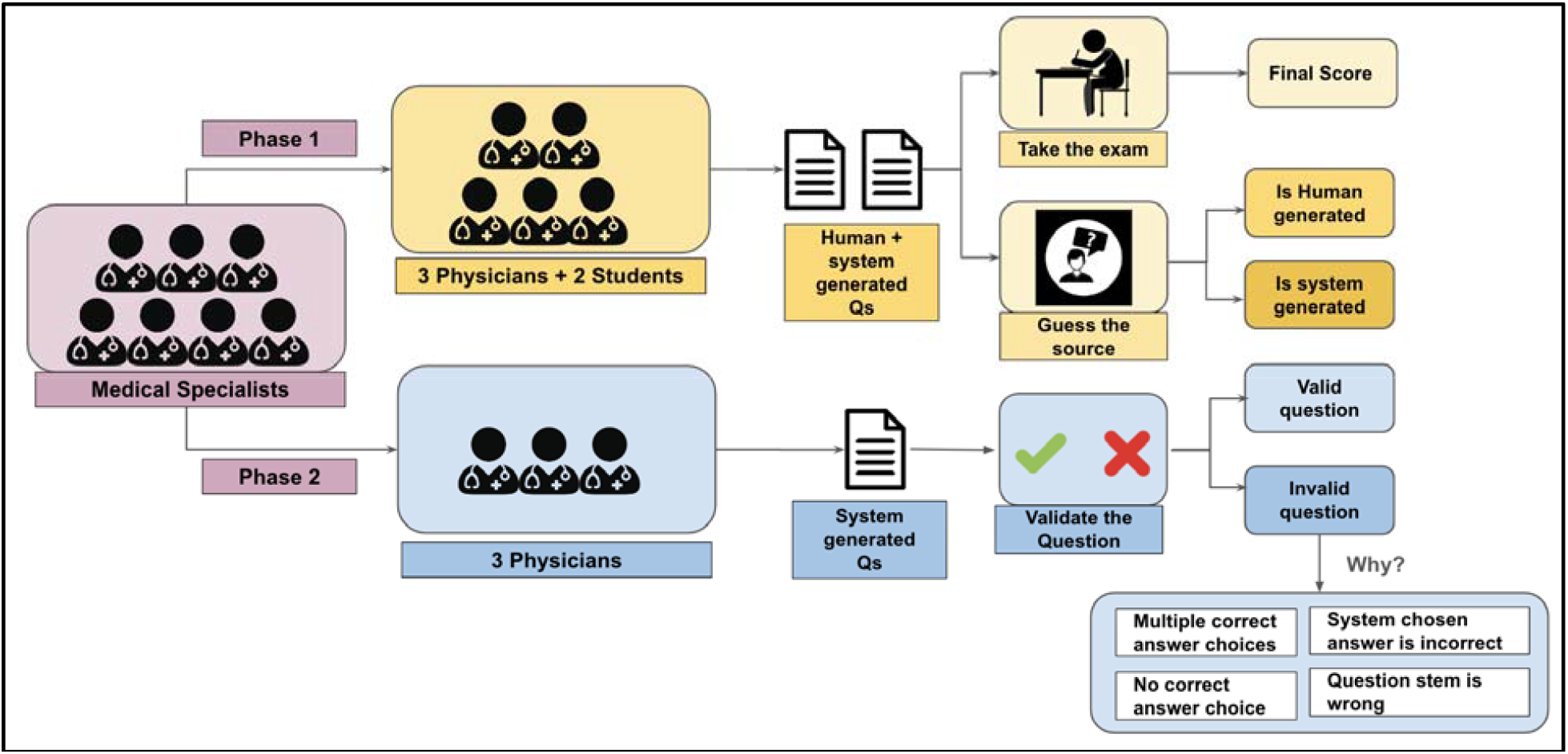
Evaluation Process by Medical Specialists: In Phase 1 of the study, three physicians and two medical students attempted a USMLE exam that included both real and system-generated questions, tasked with choosing the best answer for each question and identifying which questions were system-generated. In Phase 2, three physicians evaluated the system-generated question-answer pairs to determine their validity. For invalid questions, they categorized the issues into four types: multiple correct answer choices, no correct answer choice, the system-chosen answer choice is incorrect, or the question stem is incorrect.

### 3.3. Evaluation by LLMs

An ensemble of five LLMs from the Hugging Face hub^27^ (a public repository of models) was selected for evaluation based on the models’ performance in public open LLM leaderboards^28^ and community support: Meta-Llama-3-70B-Instruct from Meta, Mixtral-8×22B-Instruct-v0.1 from Mistral AI, Qwen2-72B-Instruct from Alibaba, Phi-3-medium-4k-Instruct from Microsoft, and llama-2-70b-chat from Meta. Each of these models brings unique strengths due to variations in their architectures and training datasets, providing diverse perspectives on the task of identifying the best answer to system-generated USMLE-style questions.

To evaluate the validity of the system-generated question-answer pairs, we tasked the ensemble with selecting the best answer. A simple majority-based classifier was constructed to flag potentially flawed questions: if any of the models disagreed with GPT-4’s selected best answer, the question was flagged for further review. This design is grounded in ensemble learning theory ^29^, which posits that combining predictions from multiple models can improve accuracy and reliability by reducing individual model bias and variance. The assumption here is that even a single disagreement may indicate a potential error, ambiguity, or inconsistency in the question or answer choices. This model heterogeneity strengthens the system’s ability to detect flaws by increasing the likelihood of identifying subtle inconsistencies.

Conversely, when all models in the ensemble agreed with GPT-4’s answer, the question was considered less likely to be flawed. This consensus-based approach is consistent with voting schemes in ensemble learning, where agreement across multiple models typically indicates a high confidence prediction. ^30^

### 3.4. Categorization and Post-Hoc Editing by GPT-4

GPT-4 was prompted to categorize each question-answer pair in the 100-item set into one of 18 categories outlined in the USMLE content outline^31^. This categorization aimed to evaluate whether the system-generated questions fell into the same categories as the original human-generated questions used in the prompts. A physician then reviewed these category assignments for each question to verify their validity. We caveat the evaluation of category assignment validity by noting that the ground truth categories per question are not made publicly available by the NBME and the team of physicians and medical students that imputed item categories have no affiliation with NBME/USMLE.

For the system-generated questions deemed incorrect by the ensemble of LLMs and physicians, we conducted a post-hoc editing stage. During this stage, we asked GPT-4 to first identify why a specific question was wrong and then modify or correct the mistakes present in the flagged questions. This was done to assess GPT-4’s capability to improve its responses based on iterative feedback and refinement. A physician manually evaluated each corrected exam item to assess whether the corrections led to a valid result.

The data collection and analysis were conducted in multiple phases from April 2023 to December 2023, and an updated evaluation analysis was completed in July 2024. This evaluation process aimed to provide insights into the practical applications of AI in generating high-quality exam questions and to assess the potential of GPT-4 in contributing to medical education. Data and code for reproducing our results can be found at https://github.com/som-shahlab/gpt4usmle.

## 4. Results

### 4.1. Evaluation by Physicians

The average accuracy of the three physicians and two medical students in distinguishing between human-generated and GPT-4-generated questions was 51.8% (95% CI: 46.1% to 57.5%), nearly equivalent to random guessing. This suggests that there were no identifiable systematic differences between the system-generated and human-generated questions. Furthermore, the Pearson correlation between participants’ performance on system-generated and human-generated questions was statistically significant (ρ = 0.952, *P*-value = 0.012). This strong correlation indicates that system-generated questions are comparable to human-generated questions in their ability to assess the clinical knowledge and skills examined in USMLE-style questions, providing a reliable and consistent measure of performance across both question types.

On a separate task where three physician reviewers were asked to validate the 50 AI generated questions, 32 (64%) questions were deemed “correct” by all reviewers, while 18 (36%) were deemed “incorrect” by at least one reviewer. The reasons for labeling exam items as “incorrect” included “Multiple correct answer choices” (n=9), “AI-chosen answer is incorrect” (n=6), and “No correct answer choice” (n=3). These findings highlight specific areas where the system-generated questions fell short and suggest areas for further refinement in the AI’s question generation capabilities.

Reviewers spent, on average, 3.21 minutes (95% CI 2.73 to 3.69) reviewing each system-generated exam item for correctness. This quick evaluation time highlights a significant potential efficiency advantage, as it is substantially faster than drafting a question from scratch, which typically involves extensive research, drafting, and revision.

### 4.2. Evaluation by LLMs

All LLMs within our LLM ensemble achieved adequate performance on the human-generated USMLE-style exam questions (see Table 1). Our proposed LLM ensemble classifier was able to discriminate between invalid system-generated questions with an Area under the Receiver-Operator Characteristic curve (AUROC) of 0.79. We considered an item to be classified by the model as “flawed” if any one of the 5 LLMs in the ensemble disagreed with GPT-4 on the best answer choice. Of the 18 system-generated question-answer pairs deemed flawed by clinician reviewers, our approach correctly flagged 15 (Recall = 15/18 = 0.83). Overall, our approach flagged 25 system-generated question-answer pairs as flawed (Precision = 15/25 = 0.60). Of the 25 system-generated questions not flagged by our approach, 22 were deemed valid by clinicians. See Table 2.

**Table 1:** Performance of LLMs in model ensemble on human- and system-generated USMLE-style questions. All models performed reasonably well (examinees typically must answer approximately 60% of items correctly to achieve a passing score on the USMLE)^32^.

| Model | % Correct on human-generated questions | % Correct on system-generated questions |
| --- | --- | --- |
| Meta-Llama-3-70b-Instruct | 0.80 | 0.80 |
| Mixtral-8x22B-Instruct-v0.1 | 0.80 | 0.78 |
| Phi-3-medium-4k-instruct | 0.76 | 0.80 |
| Qwen2-72B-Instruct | 0.80 | 0.82 |
| llama-2-70b-chat_huggingface | 0.60 | 0.62 |

**Table 2:** Confusion matrix for the LLM ensemble used to determine whether system-generated questions are potentially invalid by analyzing whether all LLMs agree with GPT-4 on the best answer (not flagged) or at least one LLM disagrees with GPT-4 on the best answer (flagged).

|  | Flagged as invalid by LLM ensemble | Not flagged by LLM ensemble |
| --- | --- | --- |
| Deemed invalid by clinician reviewers | 15 | 3 |
| Deemed valid by clinician reviewers | 10 | 22 |

### 4.3. Categorization and Post-Hoc Editing by GPT-4

For the categories assigned to each question by GPT-4, 8 questions were assigned invalid content category labels, while the remaining 92 questions were assigned appropriate labels. This outcome shows that GPT-4 generally performed well in classifying question categories, although it occasionally struggled to differentiate between Behavioral Health and Social Sciences. This challenge might be addressed by clarifying that Behavioral Health pertains to psychiatry and mental health topics, whereas Social Sciences covers medical ethics, interpersonal health, and health system quality improvement.

Additionally, 16 out of the 50 questions matched the category of their corresponding sample question. This suggests that GPT-4 introduces a degree of variability and diversity in its generated questions. Rather than merely replicating existing content, GPT-4 demonstrates the ability to create new and varied material. A breakdown of categories can be seen in the Supplementary section at - https://www.medrxiv.org/content/10.1101/2023.04.25.23288588v2

For post-hoc editing, the questions deemed incorrect by at least one reviewer were passed through GPT-4. The model was asked to classify why a question-answer pair was incorrect and then to provide a corrected version. Impressively, for 9 out of 18 questions (50%), GPT-4 identified the same reason for incorrectness as the physician reviewers. For 11 of these 18 questions (61%), GPT-4 was able to correct its original mistake, resulting in a valid exam item. This demonstrates GPT-4’s capability not only to generate questions but also to accurately diagnose issues with them and offer corrections.

## 5. Conclusion

With ever-increasing costs of medical education, medical student debt, and a looming physician shortage^33^, there is an urgent need for cost-effective and easily accessible medical exam preparation resources. We designed QUEST-AI, a first-of-its-kind system that can improve access to high-quality USMLE-style questions by using LLMs to generate candidate exam questions, flag invalid candidate items, and correct flawed exam items. While performance of the system is not perfect, clinician evaluation suggests that (1) a significant majority of exam items generated using our approach are valid; (2) candidate performance on items generated using our approach correlates strongly with performance on human-generated USMLE-style questions; and (3) our system can be used to generate exam across a variety of content categories. This offers a promising solution for decreasing the cost and time required to generate USMLE-style questions. This in turn could reduce both the costs for exam preparation materials that debt-burdened medical students face and the costs for generating new exam items that non-profit organizations like the National Board of Medical Examiners face.

## 6. Limitations

There are several important limitations to our system to consider when assessing whether it can be used in medical education.

First, with respect to our evaluation, the medical specialists who attempted to select the best answer on the evaluation set of 50 system-generated and 50 human-generated questions were not MD students (the primary audience that would benefit from such a system); they were practicing MDs who had already passed the USMLE Step 2 CK exam and DO students who would take a different but similar exam as part of their training. This was by design: we wanted to ensure that no assessor would recognize the exam items in the publicly available NBME-provided USMLE-style practice exam. Otherwise, their ability to distinguish between human- and system-generated questions would be overly optimistic. Additional study is needed to understand whether our results translate to the primary population of interest, namely MD students preparing to take the USMLE Step 2 CK exam.

Second, the clinicians who determined whether or not the system-generated exam items were valid were not expert exam writers nor were they affiliated with the NBME. It is quite possible that system-generated exam items deemed valid by our panel of clinicians would be considered invalid by NBME-employed expert exam writers, and vice versa.

Third, there was no threshold for which our LLM ensemble-based flagging system was able to correctly recall *all* the system-generated exam items deemed invalid (except for if we trivially flagged all the items as invalid). There were 3 of 18 items deemed invalid for which all 5 LLMs in the ensemble agreed with GPT-4’s best answer selection (thus the question was not flagged) but where at least one clinician deemed the overall exam item to be invalid. This suggests that, were this system to be used entirely autonomously, it could generate flawed exam items. This has important ethical implications that should be considered and potentially addressed with improved methods before releasing the tool to the broader public.

Finally, the number of system-generated questions was relatively small, with only 50 questions included in the study. While this sample size provided useful insights for an initial evaluation, it limits the generalizability of the findings. A larger set of questions is needed to provide a more comprehensive assessment of the system’s performance across different content areas and question formats. Increasing the sample size in future studies will also allow for a more detailed evaluation of additional metrics and improve the statistical power of the results.

## Data Availability

All data produced in the present study will be made available upon reasonable request to the authors

https://github.com/som-shahlab/gpt4usmle

## 8. Acknowledgment

Preprint of an article published in Pacific Symposium on Biocomputing © 2024 World Scientific Publishing Co., Singapore, http://psb.stanford.edu/.

## Supplementary Material

**eTable 1:**
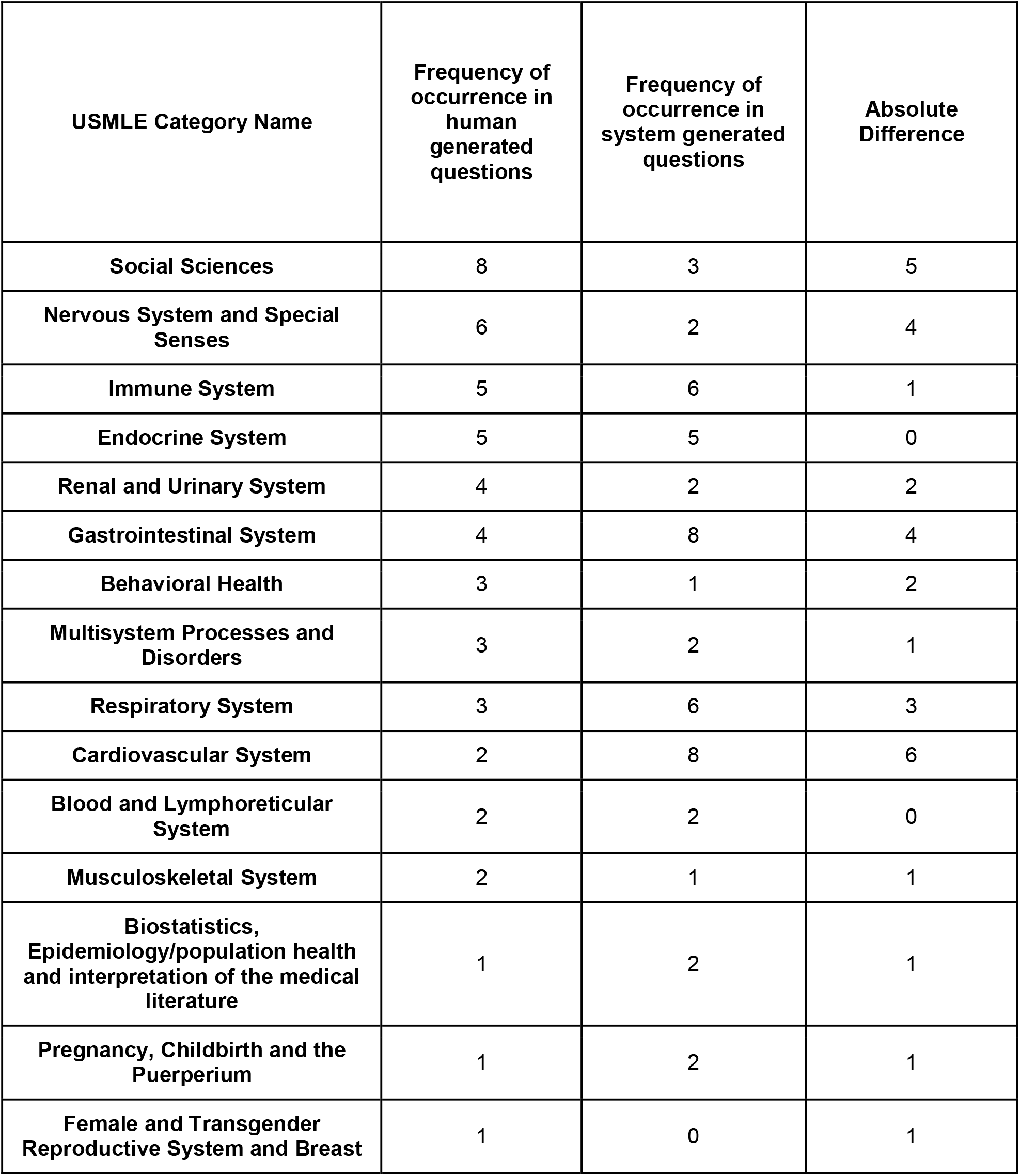
Category assignments for human- and system-generated USMLE-like questions: The first column is the USMLE category name, the second and third columns are the frequency of occurrence of each category in human and system generated questions respectively and the last column is the absolute difference between these frequencies.

